# The association of neurofibromatosis and autism symptomatology is confounded by behavioral problems

**DOI:** 10.1101/2019.12.20.19015420

**Authors:** Hadley Morotti, Sarah Mastel, Kory Keller, Rebecca A. Barnard, Trevor Hall, Brian J. O’Roak, Eric Fombonne

**Affiliations:** OHSU: Department of Molecular and Medical Genetics; OHSU: Department of Pediatrics & Institute on Development & Disability; OHSU: Department of Psychiatry

## Abstract

**Aim:** to evaluate if autism symptoms and diagnoses are raised in children with neurofibromatosis type 1 (NF1), to which levels, and to determine if co-occurring symptomatology accounts for this elevation.

**Method:** We interrogated our hospital electronic medical records. We collected parental reports of autism symptomatology, adaptive behavior, and co-occurring behavioral and emotional problems on a subsample of 45 children (9 years 2 months, 49% male). Age- and sex-matched controls with (N=180) or without ASD (N=180) were drawn from the Simons Simplex Collection and compared cross-sectionally to participants with NF1.

**Results:** Diagnoses of ADHD (8.8%), not of ASD (2.1%), were raised among 968 children with NF1 identified through electronic search. Mean Social Responsiveness Score (55.9) was below the cut-off of 60 for significant autism symptoms. Participants with NF1 had significantly more autism and behavioral symptoms than typically developing (TD) controls, and significantly less than controls with autism, with one exception: ADHD symptom levels were similar to those of autistic controls. When emotional, ADHD, and communication scores were covaried, the difference between participants with NF1 and TD controls disappeared almost entirely.

**Interpretation:** Our results do not support an association between NF1 and autism, both at the symptom and disorder levels.

**What this paper adds:**

- Diagnoses of ADHD, not of ASD, were raised among children with NF1.
- Increases in autism symptoms did not reach clinically significant thresholds.
- Co-occurring ADHD symptoms accounted for increased autism questionnaire scores.
- Adaptive behavior in NF1 participants showed normal socialization but lower communication proficiency.

## Introduction

Neurofibromatosis type 1 (NF1) is a rare (1 in 3,000; Friedman, 1999) neurocutaneous disease affecting multiple organs and caused by pathogenic variants in the NF1 gene (17q11.2). NF1 patients have a raised risk of neurodevelopmental problems such as cognitive and learning deficits, motor delays, social skills impairments, emotional dysregulation, anxiety symptoms (Torres Nupan et al., 2017; Vogel et al., 2017; Johnson et al., 1999; Pasini et al., 2012) and of attention deficit hyperactivity disorder (ADHD) symptomatology and disorders (Lidzba et al., 2012).

An increased prevalence of NF1 in epidemiological samples of children with autism spectrum disorder (ASD) diagnoses has been reported by some (Timonen-Soivio et al., 2016; Bilder et al., 2016) but not all (Fombonne et al., 1997; Mouridsen et al., 1992; Saemundsen et al., 2013) population-based studies of ASD. Given the base rate of NF1 is low, samples of ASD participants much larger than those usually surveyed are necessary to detect with confidence a prevalence increase of NF1. By contrast, relatively smaller NF1 samples are required to detect an increase in the prevalence of ASD currently estimated to be in the 1% to 2% range (Meyers et al., 2019; Baio et al., 2018). In a population-based NF1 registry, a study reported a prevalence of 24.9% for ASD (45.7% with a broader ASD definition) using standardized research diagnostic tools (Garg et al., 2013b). A study of consecutive referrals to an NF1 clinic found a minimum prevalence of 26% for formally diagnosed ASD (Plasschaert et al., 2014). Evaluating autism symptoms rather than diagnoses, other studies sampling clinical series of children with NF1 indicated that as many as 50% of individuals with NF1 may meet diagnostic criteria for ASD based on questionnaire thresholds (Garg et al., 2013a; Walsh et al., 2016; Constantino et al., 2015; Morris et al., 2016).

However, uncertainty remains regarding the nature of the association between NF1 and ASD. Specifically, raised levels of ASD symptoms may not translate into true co-occurring ASD diagnoses, and the association between NF1 and ASD may be confounded by ADHD symptoms and diagnoses, and other co-occurring behavioral problems. Prior studies have not consistently controlled for these covariates. Additionally, evaluations of ASD symptoms and diagnoses rely heavily on parental reports that have been shown in several recent studies not to discriminate well between ADHD and ASD symptoms and, in the presence of behavioral problems, to inflate ASD symptom scores on both diagnostic interviews and parent-completed questionnaires (Grzadzinski et al., 2016; Havdahl et al., 2016; Hus et al., 2013)

The specific objectives of this study were: 1) to estimate the association between ASD, ADHD and NF1 as categorical ICD-10 diagnoses; 2) to evaluate if ASD symptoms in NF1 are elevated and, if so, to determine whether ADHD and other co-occurring symptomatology accounts for this elevation.

## Method

### Participants

With the exception of controls, participants were selected from OHSU databases and specialized clinics. OHSU is the only tertiary medical center in Oregon that harbors a medical school. The clinical autism program, which is housed at OHSU’s Doernbecher Children’s Hospital, is the largest and most utilized ASD diagnostic center in Oregon. OHSU accepts patients without commercial medical insurance, which makes it a center of reference for complex childhood neurodevelopmental disorders.

#### OHSU-Electronic Medical Records (OHSU-EMR)

To address objective 1, we interrogated the electronic medical records (EMR) at OHSU, identified patients with an ICD-10 NF1 diagnosis, and examined the distribution of associated ICD-10 diagnoses of both ADHD and ASD.

#### OHSU-NF1 clinic (OHSU-NF1)

From the OHSU NF1 clinic, we recruited a sample of participants aged 4 to 12 for additional study. From 264 NF1 patients meeting age and diagnosis criteria for inclusion, 45 responded positively to a mailed invitation letter.

#### Simons Simplex collection controls with (SSC-ASD) or without (SSC-TD) ASD

Participants with or without ASD from the Simons Simplex Collection (SSC) were used as controls. The SSC has one the largest collection of trios (1 ASD proband and 2 biological parents) and quads (trio plus 1 unaffected sibling) that has contributed to substantial genetic discoveries. All families were phenotyped in depth (detailed in Fischbach and Lord, 2010) and data were released for future research in a publicly accessible repository (www.sfari.org). We searched the SSC database for probands with ASD and for unaffected siblings, who were selected independently, allowing most participants to be from different families. SSC participants were matched on age and sex to each OHSU-NF1 clinic participant in a 4:1 ratio in order to create two control groups of 180 participants with ASD and 180 typically developing (TD) unaffected siblings.

### Instruments & Data

EMR were searched for patients with an ICD-10 code of NF1. For these subjects, all co-occurring ICD codes of ASD (or Pervasive Developmental Disorders: PDD; section F84.x in ICD-10, excluding Rett syndrome) and of ADHD (F90.0 – F90.9) were obtained.

We then selected subjects with an NF1 diagnosis, aged 5 to 12 years, for a more detailed investigation. The age range was restricted due to limited resources and suitability for our research measures.

After completion of the informed consent process, parents were mailed a package of questionnaires that included

a. *Child Behavior Checklist (CBCL:* Achenbach and Rescorla, 2001): 120 items covering problem behaviors over the last 6 months, each rated on a 0-2 scale. T-scores were used for the narrow band syndrome labeled “attention deficit-hyperactive syndrome,” and the externalizing, internalizing, and total scores.
b. *Vineland Adaptive Behavior Scales, Second Edition (VABS-II;* Sparrow et al., 2005): We used the parent self-administered version. The instrument is normed and generates 3 domain scores for Communication, Daily Living Skills, and Socialization, and a total Adaptive Behavior Composite (ABC) score. All scores are standardized with a mean of 100 and a SD=15.
c. *Social Responsiveness Scale, second edition (SRS-2*; Constantino and Gruber, 2005): It includes 65 statements scored 0 to 3 on a Likert-type scale, and it yields a total raw and t-score. Subscale scores were not used. T-score over the cut-off of 60 has been suggested to indicate clinically significant ASD symptoms, and >=75 for predicting a clinical diagnosis of ASD. We analyzed the SRS-2 both dimensionally (with the T-scores) and categorically (using cut-offs of 60 and 75). In addition, for comparative purposes, we used published SRS data from the International NF1-ASD Consortium Team (INFACT; Morris et al., 2016).
d. *Repetitive Behavior Scale-Revised (RBS-R;* Bodfish et al., 1999): It contains 44 items, scored 0-3; 6 subscales and one Total score can be derived. We used the total RBS-R score for our analyses. Because it was designed to capture atypical behaviors, RBS-R was not collected on the SSC-TD controls.
e. *Social Communication Questionnaire (SCQ*; Rutter et al., 2001*), lifetime version*: It comprises 40 items scored yes=1/no=0. We used the total score and the proportion over the cut-off of 15 that identifies a broad ASD definition (Berument et al., 1999).

### Data and Statistical Analysis

Conventional statistical tests were used for categorical variables. To account for unequal sample sizes and unequal variances, mean differences across the three groups were tested with ANOVA and robust Brown-Forsythe statistics. Follow-up pairwise comparisons were performed with post-hoc Games-Howell tests. Normality assumptions were checked with Wilk-Shapiro test, and when appropriate, non-parametric tests were employed. Stepwise multiple logistic regression was used to evaluate whether or not membership to SSC-TD or OHSU-NF1 was predictive of high versus low scorers on the SRS-2 after adjusting on non-ASD covariates. Throughout, a p-value of 0.05 was retained as level for statistical significance.

### Ethical Approval

The OHSU IRB approved the study. Written consent and assent to participation in the study and publication of its results was collected from participants from the OHSU-NF1 clinic sample. Simons Simplex Collection was a study with multisite IRB approval. All participants consented to participate and to have their data stored in a repository for future research use.

## Results

### OHSU-EMR sample

In the OHSU-EMR sample, 968 patients had an NF1 diagnosis of whom 20 (2.07%; 95% CI: 1.3%-3.2%) had a co-occurring ICD-10 PDD/ASD diagnosis and 85 (8.8%; 95% CI: 7.2%-10.7%) had an ADHD diagnosis.

### OHSU-NF1 clinic sample

In the OHSU-NF1 clinic sample, the 45 NF1 participants did not differ from the 219 non-participants with respect to gender, race, ethnicity, or age (all NS). When compared to both control groups, the participants with NF1 did not differ for the matching variables age (Table 1) and sex (48.9% male in all groups), as well as for Hispanic status (overall 9.9%; χ^2^=.53, 2 df; p=.77) and race (overall 80.2% White; χ^2^= 2.91, 2 df; p=.23).

**Table 1:**
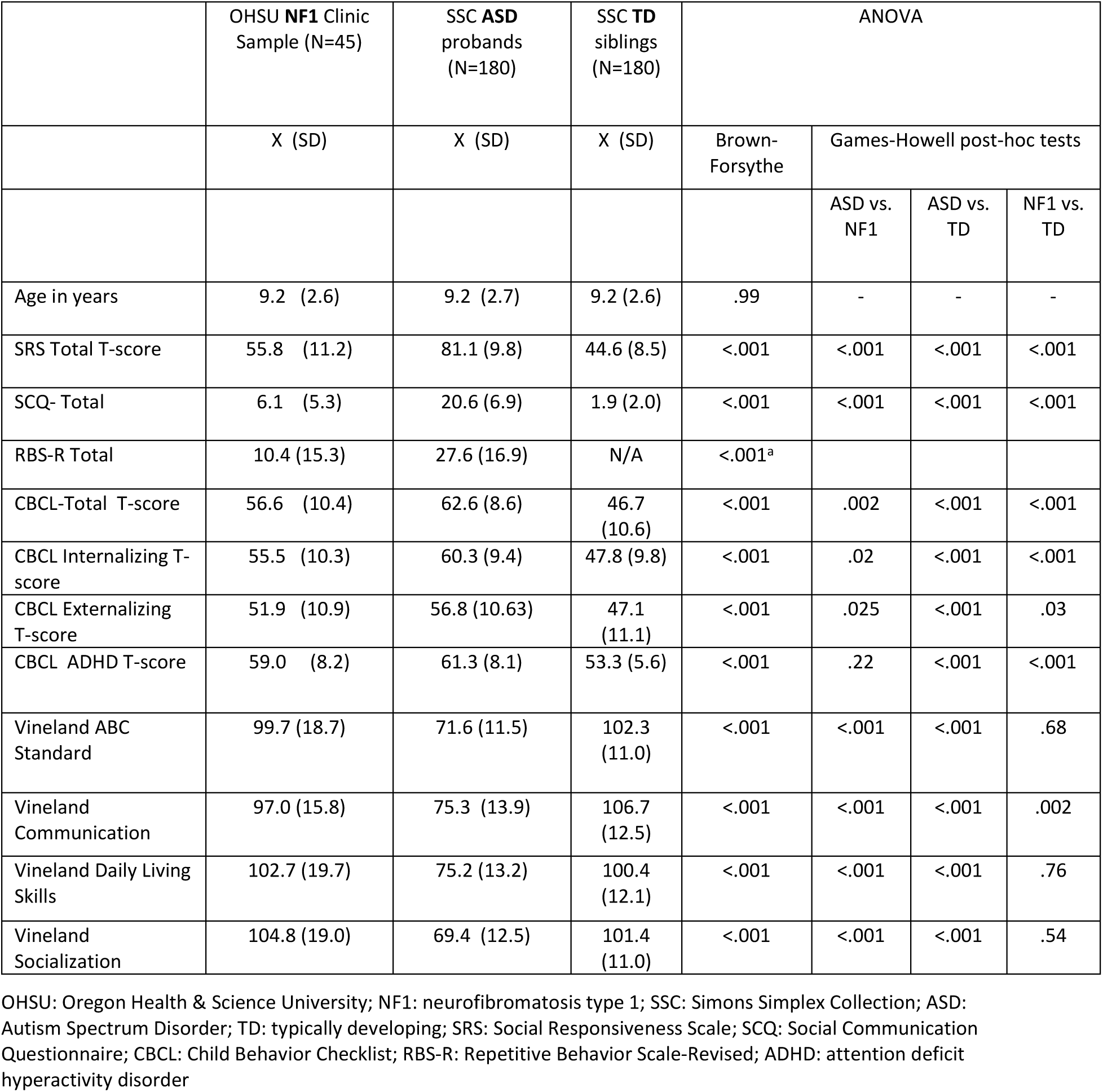
Comparison of participants with neurofibromatosis 1 with age- and gender-matched controls, with or without autism.

Significant differences were found between the three groups on measures of autistic symptomatology (SRS-2, SCQ). Post-hoc tests showed that the NF1 group scored significantly lower than the SSC-ASD group and significantly higher than SSC-TD controls (Table 1). Mean scores of the NF1 group were much closer to the SSC-TD than to the SSC-ASD controls. In addition, participants with NF1 had significantly lower scores than controls with ASD on the Repetitive Behavior Scale. Compared to the INFACT consortium data (Morris et al., 2016), the mean SRS-2 score for our NF1 sample was slightly lower (55.8 vs 58.2); remarkably, means in both studies fell below 60, within the normal range of SRS norms (Figure 1). The proportions of OHSU-NF1 participants with SRS-2 scores >=60 or >=75 were 40.0% and 6.7%, similar to INFACT consortium data (respectively, 39.2% and 13.2%; Morris et al., 2016). The three groups were also significantly different on the three broad-band mean scores of the CBCL and the narrow-band ADHD subscale (Table 1). Pairwise comparisons showed that there were significant differences for all between-pair comparisons except for the ADHD subscale where no difference was found between the NF1 and SSC-ASD groups. Significant differences were found for all Vineland scores in omnibus tests. Post-hoc comparisons showed that NF1 participants did not differ from SSC-TD participants on the Vineland ABC, Daily Living Skills, and Socialization mean scores. However, NF1 subjects had significantly lower mean scores than SSC-TDs on the Communication domain although their mean scores remained close to the population mean; the difference with SSC-ASD controls was statistically significant and of a much larger magnitude. Interestingly, NF1 and SSC-ASD subjects differed most on the Vineland Socialization domain, and the average Socialization score of the NF1 group was above the population mean and the SSC-TD control mean as well.

**Figure 1.**
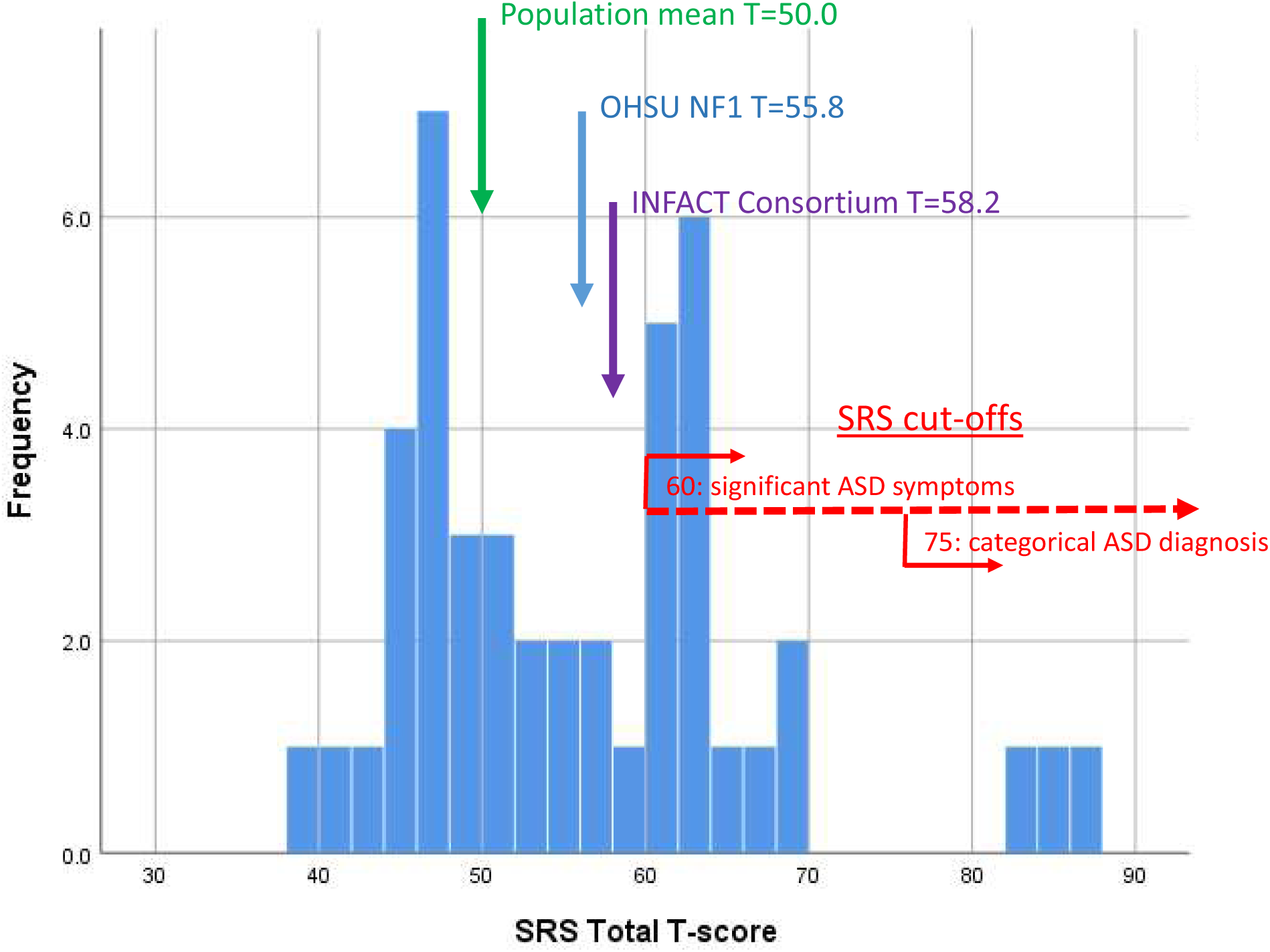
Distribution of Social Responsiveness Scale (SRS) scores in the OHSU NF1 clinic Sample. Footnote: The Figure shows the position of population mean in a normative sample (red), and the mean in the INFACT Consortium multisite sample of 531 NF1 patients (green; Morris et al., 2016) OHSU: Oregon Health & Science University; NF1: neurofibromatosis type 1; SRS: Social Responsiveness Scale; INFACT: International NF1-ASD Consortium Team

In the whole sample, correlations (Spearman’s rho) between the SRS-2 and the CBCL and Vineland scores were all significant (p<.01): CBCL Total (.79), CBCL Externalizing (.59), CBCL Internalizing (.66), CBCL ADHD (.66), Vineland Communication (-.76), Vineland DLS (-.69), Vineland Socialization (-.79), Vineland Adaptive Behavior Composite (-.79). The pattern of correlations was similar when inspected in the NF1 sample only. There was no sex difference in SRS-2 scores (t=.78; p=.43) and in any other behavioral or adaptive score (all p’s>.10); age was weakly (rho: -.10 to -.15) negatively correlated with Vineland scores only.

To address objective 2, we then compared NF1 and SSC-TD participants in order to test if the between-group difference in SRS-2 scores could be attributed to co-occurring ADHD and other behavioral problems. First, in a univariate ANOVA, 20% of the overall SRS variance was initially explained by group (SSC-TD or NF1) membership (Table 2, Model 1, **bold**). Next, when ADHD symptom score was added as covariate (Table 2, Model 2), the proportion of SRS-2 variance explained rose to 43%, with ADHD scores accounting for over 30% of it. The variance explained by group membership was reduced to 8.7% (**red bold**). Finally, in a fully adjusted model (Model 3), group membership accounted only for 3.2% of the SRS-2 variance, much less than that explained by the CBCL-ADHD scale, the CBCL Internalization score and the Vineland Communication standard score, each taken alone or in combination. Among independent variables, CBCL-ADHD scores alone accounted for the highest proportion of SRS-2 variance. Between Models 1 and 3, the SSC-TD vs NF1 difference in estimated marginal means was reduced from 11.2 to 3.4, a difference that is clinically negligible.

**Table 2.** SSC-TD-NF1 difference in SRS scores is accounted for by co-occurring behavioral problems - ANCOVAs models (N=224)

| | Estimated marginal means | | Mean difference (95% CI) | | | $p$ | F | $p$ | Partial Eta Squared |
| --- | --- | --- | --- | --- | --- | --- | --- | --- | --- |
|  | TD | NF1 | Mean (SE) | Lower | Upper |  |  |  |  |
| **Model 1** |  |  |  |  |  |  |  |  |  |
| SSC-TD/NF1 | 44.58 | 55.76 | 11.18 | 8.19 | 14.17 | <.001 | 54.3 | .000 | **.197** |
|  |  |  |  |  |  |  | 54.3 | .000 | **.197** |
| **Model 2** |  |  |  |  |  |  |  |  |  |
|  | 45.56 | 51.84 | 6.27<br>(1.38) | 3.55 | 8.99 | <.001 | 83.5 | .000 | **.435** |
| SSC-TD/NF1 |  |  |  |  |  |  | 20.6 | .000 | **.087** |
| ADHD T score |  |  |  |  |  |  | 94.1 | .000 | .303 |
| **Model 3** |  |  |  |  |  |  |  |  |  |
|  | 45.82 | 49.25 | 3.43<br>(1.32) | 0.84 | 6.03 | .010 | 56.9 | .000 | **.521** |
| SSC-TD/NF1 |  |  |  |  |  |  | 6.8 | .010 | **.032** |
| CBCL ADHD T score |  |  |  |  |  |  | 37.9 | .000 | .153 |
| CBCL_Internalizing T score |  |  |  |  |  |  | 26.5 | .000 | .113 |
| Vineland Communication Standard score |  |  |  |  |  |  | 15.8 | .000 | .070 |

NF1: neurofibromatosis type 1; SSC: Simons Simplex Collection; TD: typically developing; SRS: Social Responsiveness Scale; SCQ: Social Communication Questionnaire; CBCL: Child Behavior Checklist; RBS-R: Repetitive Behavior Scale-Revised; ADHD: attention deficit hyperactivity disorder

Because it was possible that the relationship between ADHD and SRS-2 symptoms might not be uniform along the continuum of SRS-2 scores, we dichotomized the SRS-2 using a T score of 60 or higher. This created two groups of participants, with 28 subjects (18 NF1, 10 SSC-TD) designated as high SRS-2 scorers. We ran a stepwise multiple logistic regression analysis with age, sex, group (NF1/SSC-TD), all CBCL and Vineland scores. The final model retained CBCL-ADHD score (OR=1.14; 95%CI: 1.05-1.23), Vineland Communication standard score (OR=.93; 95%CI:.88-.97) and CBCL-Internalizing score (OR=1.07; 95% CI:1.01-1.14). Group never entered the model. The model had good fit with the data (Hosmer and Lemeshow χ^2^: p=.63).

## Discussion

This study has several strengths and limitations. Limitations include a small sample size, a low participation rate, reliance on parental reports, and the absence of longitudinal data. We did not have data on the NF1 medical and cognitive severity or on characteristics of the mutations. Our sample included school-age children only, thus extrapolation of findings to adolescent and adult samples is unwarranted. Among the strengths, were the examination of ASD both at a symptom and diagnostic levels, the recruitment of two well characterized control groups with large sample sizes that provided adequate statistical power, and statistical comparisons that were adjusted on multiple covariates.

The results of our study do not support an association between NF1 and autism defined as either a diagnosable disorder or raised level of autistic symptomatology. Compared to generally accepted population estimates of 1.7% for ASD (Baio et al., 2018) and 5% for ADHD (Cortese and Coghill, 2018), the prevalence of these diagnoses (2.1% and 8.8%, respectively) in the OHSU EMR sample, taken in conjunction with their associated confidence intervals, suggested no association of NF1 with ASD, but a significant association with ADHD at a diagnostic level. The increase in ADHD diagnoses is in agreement with most previous studies of children with NF1 that have reported even higher rates (e.g. 38.3% in Hyman et al., 2005). As indicated earlier, our findings for ASD diagnoses are discrepant with some recent studies. Thus, Garg et al. (2013a & 2013b) reported a prevalence of 45.7% of broad ASD and 24.9% for a narrower ASD definition in a registry study of 109 children ages 4 to 16 with NF1. Several aspects of that investigation need to be borne in mind. Response rate was low (52.7%) for the screening phase and despite properly weighting the data to account for varying selection and participation rates at different phases, no behavioral data on non-responders were available to rule out a selection bias. Thus, if individuals with both NF1 and ASD were more likely to participate, this could account for at least part of the findings. In addition, ASD diagnoses were mechanically derived from algorithms based on IQ, ADOS and ADI scores but without infusion of clinical judgment. The fact that 26 of 32 (81.2%) of children diagnosed with ASD also had an ADHD diagnosis would require a more clinically informed interpretation of overlapping symptomatology. It is also noteworthy that only two children in this sample had previously received a diagnosis of ASD, a 1.9% prevalence that is much lower than the survey estimate of computer generated diagnoses; moreover, teachers’ reports were at variance with parental ones (5.7% vs 29.4% SRS score in ASD diagnosis range; Garg et al., 2013a). Finally, correlations between measures of ADHD and ASD symptoms were not reported and other aspects of the study (low male: female ratio, low reliability on social deficits measures) called for a cautious interpretation of the results. Similarly, the study by Bilder et al. (2016) identified a prevalence of 0.18% of NF1 in a large population-based sample (N=12,271) of children age 8 surveyed by CDC, a four-fold increase compared to population estimates for NF1. Of the 22 children with both NF1 and ASD, 23% had an ADHD diagnosis and only half had a previous ASD diagnosis in their record. As discussed elsewhere (Fombonne, 2018), the CDC methodology has several limitations including samples that are not representative and case definitions that are based on record reviews and diagnostic rules (e.g. having school eligibility for ASD services is equated to a confirmed ASD diagnosis) that have uncertain validity.

Most other studies have relied on questionnaire definitions of ASD and parental reports to collect information on autistic symptomatology and co-occurring problems. In the largest study, the INFACT Consortium recently investigated a large sample of 531 NF1 patients recruited at six sites (Morris et al., 2016). Findings suggested a shift of the SRS-2 score distribution toward the abnormal range (39.2% with SRS-2 >=60), and a substantial correlation between ADHD and autistic symptom scores (.61), results that parallel ours. No diagnostic confirmation of ASD was available, ADHD scores were missing for 61% of participants, and measurement confound by other co-occurring problems (e.g. anxiety) could not be evaluated due to lack of other behavioral measures. Further analyses of the ADHD-ASD symptom relationship suggested that SRS-2 scores were much higher in the high-ADHD than in the low-ADHD subgroups. Other questionnaire-based studies have reported similar elevations of autism scores among NF1 samples together with raised levels of ADHD symptomatology, and substantial correlations between these two symptom domains (Constantino et al. 2015; Walsh et al., 2012; Garg et al., 2013a). However, lack of proper control groups and of measures of comorbid problems other than ADHD leave these descriptive results subject to further interpretation.

To the best of our knowledge, this study is the first to empirically assess the extent to which raised autistic symptomatology among NF1 participants is attributable to both co-occurring ADHD and emotional symptoms. Specifically, analyses of covariance confirmed that the differences between NF1 and TD participants in levels of autism symptomatology were almost entirely accounted for by co-occurring ADHD and emotional problems. Furthermore, the NF1 participants have Socialization Vineland scores at or above the population mean in sharp contrast with published literature which shows consistent troughs with that Vineland domain in ASD samples. Our findings are consistent with new research indicating that ASD symptom measures cannot be interpreted without taking into account non-ASD co-occurring conditions. For example, Havdahl et al. (2016) clearly demonstrated that parent reports of autism symptoms on the SRS were raised for children with emotional/behavioral problems without ASD to levels comparable to those of children with ASD but without co-occurring problems. Similarly, in a study of verbally fluent school-age children diagnosed with either ASD or ADHD, Grzadzinski et al. (2016) showed that the Autism Diagnostic Interview-Revised, a gold-standard diagnostic measure based on parental interviewing, lost its discriminant power in contrast to the ADOS, a clinician based assessment, suggesting that parent reporting of social difficulties may be non-specific to autism. Examination of a large sample of subjects with ASD and their unaffected siblings also demonstrated that, in both groups, scores on the SRS increased in the presence of behavioral problems as well as cognitive and expressive language limitations (Hus et al., 2013). Authors cautioned about interpreting raised SRS scores as specific indicators of autism severity or social deficits and suggested that they rather reflect general impairment levels. Our results concur with and support that interpretation. Rather than rely on a single parent report questionnaire or even interview, investigators should adhere to the recommended multiple informant approach and, when assessing autism in the context of associated psychopathology and developmental problems, they should keep clinical judgement as the main interpretative tool. Also, and in line with our skeptical review and own findings, we note that recent guidelines from the American College of Medical Genetics and Genomics on the management of adults with NF1 do mention cognitive impairments and ADHD symptomatology but not autism (Stewart et al., 2018). This is inconsistent with claims of a raised ASD prevalence in childhood to levels of 25-50% since autism does not disappear in adult life. Nevertheless, the raised scores on the SRS and other behavioral measures point at the broad spectrum of cognitive, executive function, attention, language, and emotional processing that has long been described among patients with NF1.

## Data Availability

The original data collected at OHSU are stored on site and available upon request.
The SSC data are available through application to SFARI Base.

https://base.sfari.org

## Acknowledgements

We gratefully acknowledge funding from the Edna S. Green Trust and the OHSU School of Medicine. We thank the clinicians from OHSU’s Molecular & Medical Genetics Department, Dr. Paul Barnes’s participation to our research team, the CDRC Genetics Clinic, and the Institute on Development and Disability for their assistance in recruitment. Finally, we deeply appreciate the time and effort of all participating families.

We are grateful to all of the families at the participating Simons Simplex Collection (SSC) sites, as well as the principal investigators (A. Beaudet, R. Bernier, J. Constantino, E. Cook, E. Fombonne, D. Geschwind, R. Goin-Kochel, E. Hanson, D. Grice, A. Klin, D. Ledbetter, C. Lord, C. Martin, D. Martin, R. Maxim, J. Miles, O. Ousley, K. Pelphrey, B. Peterson, J. Piggot, C. Saulnier, M. State, W. Stone, J. Sutcliffe, C. Walsh, Z. Warren, E. Wijsman).

We appreciate obtaining access to phenotypic data on SFARI Base. Approved researchers can obtain the SSC population dataset described in this study by applying at https://base.sfari.org.

## Abbreviation List

NF1: neurofibromatosis type 1
ADHD: attention deficit hyperactivity disorder
CBCL: Child Behavior Checklist
RBS-R: Repetitive Behavior Scale-Revised
SRS-2: Social Responsiveness Scale-2^nd^ edition
SCQ: Social Communication Questionnaire
ASD: Autism Spectrum Disorder
PDD: Pervasive Developmental Disorder

